# Survival Analysis of Time to First Antenatal Care Visit among Pregnant Women in Ethiopia: Application of Parametric and Non-Parametric Model

**DOI:** 10.1101/2024.12.14.24318967

**Authors:** Yitagesu Eshetu Ayele, Zelalem Tolosa Gada, Tigist Getachew Addisu

## Abstract

**Objective:** Antenatal care (ANC) involves monitoring pregnant women to ensure the health of both mother and fetus. This study aimed to assess the timing of the initial ANC visit and its determining factors among Ethiopian pregnant women.

**Material and Methods:** The analysis used data from the 2016 Ethiopian Demographic and Health Survey (EDHS), focusing on women aged 15-49 who had a live birth within five years prior to the survey across nine regional states and two city administrations. Descriptive statistics and parametric shared frailty models were applied to identify factors influencing the timing of the first ANC visit.

**Results:** Among 7,559 women, only 20.4% attended their first ANC visit within the recommended 12 weeks. The median time to the first ANC visit in Ethiopia was 18 weeks (4.5 months). Factors that delayed the first ANC visit included rural residence (□= 1.29), higher birth order (□= 1.06-1.21), and long distance to a health facility (□= 1.08). Conversely, factors that shortened the time to the first ANC visit included higher levels of maternal education (□= 0.87-0.91), higher wealth index (□= 0.86-0.87), media access (□= 0.92), and involvement in healthcare decisions (□< 1).

**Conclusion:** Rural residence, higher birth order, and long travel distances delay the first ANC visit, while higher education, urban residence, media access, and greater wealth accelerate it. The study recommends enhanced community education on the importance of early ANC visits through health extension workers and stakeholders.

## Introduction

The World Health Organization (WHO) defines antenatal care as critical health care related to pregnancy that can be provided at home or in a medical facility. It is an important component of mother and child health [1]. According to WHO recommendations, women whose pregnancies are proceeding normally should have at least four ANC visits with individualized visit intervals; the first scheduled in the first trimester should occur before 16 weeks; Visits one, three, and four are scheduled for 24-26 weeks, 28-32 weeks, and 36-38 weeks [2].

The World Health Organization estimates that between 1990 and 2015, maternal mortality from unwanted pregnancy and difficulties associated to childbirth accounted for 303,000 maternal deaths, which is still a very high rate [3]; additionally, 2.6 million babies were stillborn globally in 2015, and 2.7 million babies died during their first 28 days of life, according to estimates of stillbirth rates from national, regional, and global sources. [4], [3]. In 2015, developing nations accounted for almost 99% (302,000) of all maternal deaths worldwide, with sub-Saharan Africa accounting for 66% (201,000) of these deaths. Approximately 830 women died each day from pregnancy- and childbirth-related preventable causes. In low- and middle-income nations, nearly all maternal deaths (99%) and child deaths (98%) took place. Despite this reduction, developing nations accounted for 239 deaths per 100,000 live births globally, compared to 12 deaths per 100,000 live births in wealthy nations (216 deaths per 100,000 live births). Ethiopia is one of ten developing nations that together account for 59% of the maternal mortality rate worldwide in 2015, with 353 deaths per 100,000 live births. Sub-Saharan Africa has the highest MMR among developing regions, at 546 deaths per 100,000 live births [3].

ANC is significantly associated with a decrease in maternal mortality and is often acknowledged as the most significant factor determining pregnancy outcomes [5]. However, only 86% and 62% of women globally receive ANC services at least once, and four or more times, respectively, during their pregnancy; even fewer pregnant women received at least four ANC visits (52% and 46% of respectively) in regions with the highest rates of maternal mortality, such as sub-Saharan Africa and South Asia [6]. Only 62% and 32%, respectively, of the women aged 15 to 49 who gave birth within the five years before to the study had at least one and four antenatal care visits (ANC) from skilled healthcare professionals, while 37% of women had no ANC visit. These rates are lower in Ethiopia than the three global average rates [7].

Unfavorable pregnancy outcomes are significantly affected by delayed ANC initiation. Evidence from Ethiopia, however, indicates that the majority of expectant mothers did not begin their first visit before the 12-week mark, as recommended by the WHO [8],[9] only 20% of women start ANC visit in the first trimester [7]. This study aimed to determine the factors influencing the time of a pregnant woman’s first prenatal care visit in Ethiopia as well as the timing of that visit.

The parameters influencing the time to the first birth after marriage were identified by the study using the parametric gamma shared frailty model. Additionally, Weibull, log-logistic, and log-normal baseline distributions were used to construct accelerated failure time (AFT) models in order to compare and determine which model best fits the time-to-first birth data. Time to first ANC visit was divided by area in this study. Therefore, the frailty term was added to the survival model in order to evaluate the region’s impact. The study’s method for identifying the variables influencing the timing of the first ANC visit during pregnancy was the parametric log logistic gamma shared frailty model.

## Material and Methods

### Data Source

Ethiopia Demographic and Health Survey (EDHS) 2016 data were used for this analysis. Using a nationally representative sample that yields estimates at the national and regional levels as well as for urban and rural areas, the Central Statistical Agency (CSA) carried out the survey from January 18, 2016, to June 27, 2016, under the direction of the Ministry of Health. The main goal of the EDHS is to provide comprehensive data on family planning, nutrition, growing kids, maternal mortality, fertility, sexual activity, and other sexually transmitted infections to decision-makers in government and planning.

### Study population

The study population included all women between the ages of 15 and 49 who were either pregnant at the time of the survey or had given birth within the five years prior to the survey (EDHS 2016). Eventually, the study included the 7559 scenarios that were weighted.

### Sampling Design

A two-stage cluster design was used to select the 2016 EDHS sample, with the census numeration area (EA) serving as the sampling unit in the first phase. The sample comprised 645 EAs, of which 443 lived in rural and 202 in urban regions. A household is included in the second sample phase. In each of the 645 EAs that were chosen, a comprehensive list of households was completed. For the 2016 EDHS, a sample of 18,008 households was chosen at random. 15,683 women participated in the full interview process, resulting in a 95% response rate [7].

### Inclusion and Exclusion Criteria

Women who are currently pregnant or have an unknown pregnancy age at the time of the survey are not eligible to participate in this study; the censoring time is calculated as the date of the pregnancy’s delivery. Women who are 15-49 years old and who gave birth within the five years prior to the survey are the only women whose eligibility criteria are met.

### Variables under the study

The response and predictor variables used in the study are defined as follows.

### Dependent Variable

The outcome of interest in this study was the time from women date of pregnant to first ANC visit which was measured in month as 0 for woman does not have ANC visit (Censored) 1for women have ANC visit (Event).

### Independent Variables

The choice of the explanatory variable include in this study are guided by literatures. Several predictor variables are considered in these studies to investigate the determinant factors for the gestational age of first ANC visit of pregnant women. covariate used in the current study are age group, residence, mothers’ education, mother’s work status, mother’s current marital status, sex of head of household, birth order of respondent, religion of respondent, wealth index of the household, whether the pregnancy was wanted/unwanted, women’s exposure to media access, woman’s involvement on health care decision, distance to health facility, husbands educational level, region.

### Methods of Survival Analysis

Survival analysis is a statistical method for data analysis where the outcome variable is the time to the occurrence of an event [10]. There are 4 main methodological considerations in the analysis of time to event data. It is important to have a clear definition of the target event, the time origin, the time scale, and to describe how participants will exit the study. The purpose of survival analysis is to model the underlying distribution of the failure time variable and to assess the dependence of the failure time variable on covariates.

### The Cox-proportional hazards model

The Cox proportional model is the most commonly used multivariable approach for analyzing survival data in medical research. It is essentially a time-to-event regression model, which describes the relation between the event incidence, as expressed by the hazard function, and asset of covariates [11].

The Cox model is written as follows:

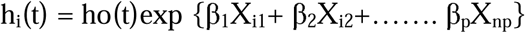

Where, ho(t) is baseline hazard, t is the failure time, X_i_ is a column vector of measured covariates for ith individual which are expected to affect the survival probability. β_1_, β_2_… β_p_ is column vector of p regression parameters. i=1, 2…. n. where n is total number of observations in the study.

### Accelerated Failure Time (AFT) Model

AFT is a parametric model that provides the alternative to the PH model whereas PH assumes the effect of covariates is to multiply the hazard by some constant, an AFT model assumes that the effect of a covariates is to accelerate or decelerate the life course of an event by some constant. In which the time to event is assumed to be a function of explanatory variables. If the proportionality assumption is not valid, the Cox proportional hazard models cannot be used in modeling, rather some parametric approaches are appropriate.

Suppose we have the values of explanatory variables recorded for each individual in a study. According to the general AFT model, the hazard function of the *i^t□^* individual is then,

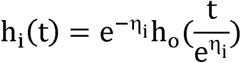

where □o(*t*) is the baseline hazard function, *ηi* is an ‘acceleration factor’ that is a ratio of survival times corresponding to any fixed value of □*it* and the acceleration factor is given according to the formula where 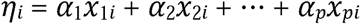.

According to the relationship of survival function and hazard function, the survival function for an individual with *p* covariates is given by

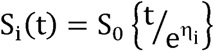

Where 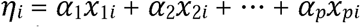 is the linear component of the model in which *x_ji_* is the value of *j*^th^ explanatory variable *X_j_* for the *it*□ individual.

### Weibull Accelerated Failure Time Model

Suppose that survival times are assumed to have a Weibull distribution with scale parameter *λ* and shape parameter *γ*, written *W (γ, λ)*, so that the baseline hazard function is

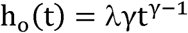

Using the result 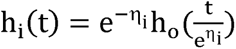, the hazard function for the i^th^ individual is then, 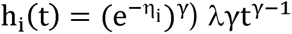

### Log-logistic Accelerated Failure Time model

One limitation of the Weibull hazard is that it is a monotonic function of time. However, situations in which the hazard function changes direction can arise.

The log-logistic distribution of the survival and hazard function are given by

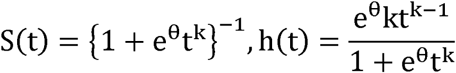

When *k* ≤ 1, the hazard function decreases monotonically and when *k* > 1, it increases from zero to a maximum and then decreases to zero.

### Log-normal Accelerated Failure Time model

The lognormal distribution is also defined for random variables that take positive values and so may be used as a model for survival data. A random variable, T, is said to have a log-normal distribution with parameters µ and *σ*, if log *T* has a normal distribution with µ and variance σ^2^

If the survival times are assumed to have a log-normal distribution, the baseline survival function and hazard function are given by

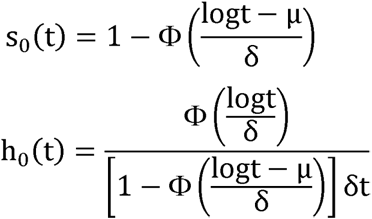

where Φ(.) is the standard normal distribution function given by

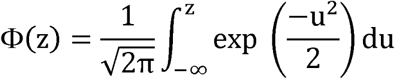

### Shared frailty models

The gamma distribution is very-well known and has simple densities. It has been widely used in parametric modeling of intra-cluster dependent because of its simple interpretation, flexibility and mathematical tractability. It is the most common distribution used for describing frailty. Even though gamma models have closed form expressions for survival and hazard functions, from a computational point of view, it fits very well to frailty data and it is easy to derive the closed form expressions for unconditional survival and hazard functions.

The survival and hazard functions of the gamma distribution are given by:

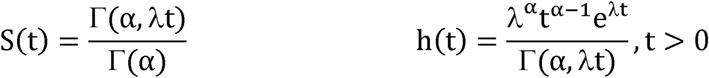

The conditional survival and hazard function of the gamma frailty distribution is given by [12].

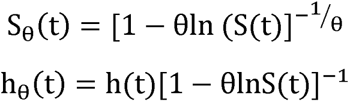

Where S(t) and h(t) are the survival and the hazard functions of the baseline distributions.

## Results

Table 1 shows the distribution of explanatory variables over all of the women at risk in the median gestational age ranging from 1 to 9 months. The Table displays the number of study participants by category, including those who had an ANC visit (event), those who had not (censored), and the percentage of women who had their first ANC visit within the first trimester (within 12 weeks).

**Table 1.** Summary of descriptive statistics for first ANC visit in Ethiopia.

| Variable | Categories | Time to first ANC visit |  |
| --- | --- | --- | --- |
|  |  | Censored No (%) | Event No (%) |
| Mother's age at birth | 15-19 | 88 (26) | 251 (74) |
|  | 20-24 | 480(33) | 974(67) |
|  | 25-34 | 1330(35) | 2489(65) |
|  | 35-49 | 920(47) | 1027(53) |
| Place of residence | Urban | 94(9.7) | 874(90.3) |
|  | Rural | 2724(41.3) | 3867(58.7) |
| Mothers Educational level | No education | 2211(78.5) | 2555(53.9) |
|  | Primary | 572 (20.3) | 1572 (33.2) |
|  | Secondary & above | 35 (1.2) | 614 (12.9) |
| Mother's work status | No | 1657(58.8) | 2404(41.2) |
|  | Yes | 1161(33.4) | 2337(66.6) |
| Current Marital Status | Never in union | 16(28.6) | 40(71.4) |
|  | Currently in union | 2628(37.1) | 4450(62.9) |
|  | Formerly in union | 174(40.9) | 251(59.1) |
| Sex of head of household | Male | 2397(37.2) | 4050(62.8) |
|  | Female | 421(37.9) | 691(62.1) |
| Religion | Orthodox | 852(29.7) | 2013(70.3) |
|  | Protestant | 632(36.8) | 1084(63.2) |
|  | Muslim | 1251(44.4) | 1566(55.6) |
|  | others | 83(51.6) | 78(48.4) |
| Wealth index of the family | Poor | 1576(47.8) | 1718(52.2) |
|  | Middle | 592(37.6) | 984(62.4) |
|  | Rich | 650(24.2) | 2039(75.8) |
| Birth order | 1 | 312(21.9) | 1112(78.1) |
|  | 2-3 | 741(44.7) | 1535(55.3) |
|  | 4-5 | 716(40.1) | 1032(59) |
|  | 6+ | 1049(49.7) | 1062(50.3) |
| Pregnancy | Wanted then | 1952(35.1) | 3605(64.9) |
|  | Wanted later | 495(37.6) | 820(62.4) |
|  | Not at all | 371(54.1) | 316(45.9) |
| Women's exposure to Media access | No | 2225(45) | 2722(55) |
|  | Yes | 593(22.7) | 2019(77.3) |
| Woman's on health care decision | No | 685(49.1) | 711(50.9) |
|  | Yes | 2133(34.6) | 4030(65.4) |
| Distance to health facility | Not big problem | 813(25.6) | 2359(74.4) |
|  | Big problem | 2005(45.7) | 2382(54.3) |
| Husband's Educational level | No education | 1705(46.9) | 1939(53.1) |
|  | Primary | 968(33.6) | 1914(66.4) |
|  | Secondary& above | 145(14) | 888(86) |
| Region/ city administrative | Tigray | 52(9.8) | 480(90.2) |
|  | Afar | 34(47.9) | 37(52.1) |
|  | Amhara | 529(32.7) | 1091(67.3) |
|  | Oromia | 1522(48.7) | 1601(51.3) |
|  | SNNPR | 486 (30.5) | 1110 (69.54) |
|  | Somali | 150(56) | 118(44) |
|  | Benishangul | 25(30.9) | 56(69.1) |
|  | Gambela | 6(28.6) | 15(71.4) |
|  | Harari | 4(23.5) | 13(76.5) |
|  | Addis Ababa | 6(3) | 191(97) |
|  | Dire Dawa | 4(12.2) | 29(87.8) |

Of the total number of women, 4741 (62.7%) had at least one antenatal care visit (ANC) from a qualified healthcare provider, whereas 2818 (37.3%) of them had none until the completion of the interview. Furthermore, just 20.4% of those expecting mothers had a visit during the first trimester.

From mother’s age at birth point of view, 51.0% were age 25-34. The proportion of ANC service started in first trimester is larger for (25-34) age groups than other age groups. Among all women 6591(87.0%) were lived in rural areas and out of this 44.0% of women’s lived urban resident start ANC visit in the first trimester while 17.0% of women’s lived in rural residents start first

ANC visit in the first trimester. More than half of the women (63.0%) have no education, (28.0%) had primary education and the remaining (9.0%) had attained secondary & higher education and 15.0% of none educated women start their ANC visit in the first trimester. Moreover, among the total women, 44.0% of the household’s wealth indexes were classified as poor while 21.0% had medium income and 35.0% were rich. 15.0% of women are from poor, while 19.0% of from medium income and 28.0% from rich income household respectively start ANC visit in the first trimester.

From household gender point of view about 85.0% of the households were male headed. Furthermore, 38.0% of the women were Orthodox, 23.0% of Protestant, 37.0% of Muslim and 2.0% of them were from other religion followers. 28.0% of orthodox follower, while 13.0% of Protestant, 18.0% of the Muslim follower start first ANC visits within WHO recommended gestational age. The result shows that, 19.0% of a women had 1st birth order, 30.0% of a women had (2-3) birth order, while 23.0% of the women had (4-5) birth orders and 28.0% of the women had six and higher birth order. The proportion of mothers using ANC service in first trimester for the first birth is relatively higher 30.0% than subsequent birth orders but only 12.0% for six and higher birth order.

According to Table 1, almost 7.0% of them were from Tigray, 0.9% from Afar, 22.0% from Amhara, 41.0% from Oromia, 1.0% from Benishangul Gumez, 21.0% from SNNPR, 2.0% from Harari, and 2.5% were from Addis Ababa, the capital. About 61.0% of mothers in Addis Ababa and Dire Dawa started visiting an ANC doctor in the first trimester, compared to only 14.0% of mothers in Somalia and 15.0% of mothers in Oromia and the SNNPR religion who saw an ANC doctor within three months.

While 46.0% of women had a job, nearly half (54.0%) did not have one. Regarding ANC visits, 24.0% of women with jobs and 18% of women without jobs received their first ANC visit during the first trimester.

The data also reveals that 4387 (58.0%) pregnant women said that travel time to the health facility was a major barrier to receiving ANC services; as a result, only 16.0% of pregnant women began their first ANC visit in the first trimester. 65.0% of the female respondents indicated they had no access to any media in terms of mass media exposure. When it came to the first trimester, just 16.0% of women without access to media began their visit, whereas 29.0% of women with media access began in the same trimester.

Only 14.0% of women who were not involved in health care decisions began their ANC visits in the first trimester, compared to 82.0% of women who were involved in health care decisions and 22.0% of women who were involved in health care decisions.

Based on the results, women’s husbands’ educational attainment, approximately 48.0% had no education, 38.0% had just completed elementary school, and 14.0% had attended secondary and higher education. Additionally, 17.0% of women without formal education begin their ANC visit in the first trimester, compared to 20.0% of mothers with primary education and 35.0% of mothers with secondary education and above.

### Non-Parametric Survival Analysis Results

#### The Kaplan-Meier estimate of Time-to-First ANC visit

Pregnant women living in rural areas had a median survival time of 6 months (95% CI: 5.8, 6.2) for the time before their first ANC visit, compared to 4 months (95% CI: [3.9,4.1]) for those living in urban areas. Women who had access to media had a longer median survival time 5 months than non-users when it came to the time until their first ANC visit. Women who lived far away from a health facility had a median survival period of six months, which was longer than that of women who lived close to one (4 months). For women without formal education, the median period of their first ANC visit was six months, longer than that of women with elementary education (4 months) and secondary education and above (3 months).

The overall median survival time of first ANC visit for Ethiopian pregnant women is 4.5 months with 95% CI (4.1, 4.9) of first ANC visit was 6 month and 3 months respectively as shown in Table 2.

**Table 2.** Summary of response variable for first ANC visit in month.

| Variable | Minimum | Maximum | Median | Mean | S.D. |
| --- | --- | --- | --- | --- | --- |
| First ANC Visit | 1 | 9 | 4.5 | 5.81 | 1.66 |

S.D.= standard deviation

#### The K-M plots of Survival and hazard functions of First ANC visit

Plots of the KM curves to the survival and hazard experience of time-to first ANC visit is shown in Fig 1. The survival plot is almost constant at beginning up to two months and decreases at decreasing rate letter. This implies that most of the women didn’t take their first ANC visit within two months during pregnancy.

#### Test of PH assumption using Schoenfeld residuals

According to [13], one technique for verifying the PH assumption is the Schoenfeld residual. According to Table 3 results, the p-value (p-value=0.000) and Chi-square value (182.04) are both significant. This suggested that there can be data that contradicts the PH assumption. The violation of the proportional hazard assumption for the covariates is also demonstrated by the scatter plots of the Scaled Schoenfeld residuals in Fig 2, which reveal that the curve was slightly upward.

**Table 3.** Test of Proportional hazard assumption by using Schoenfeld residual.

|  | chi2 | df | Prob>chi2 |
| --- | --- | --- | --- |
| **global test** | 182.04 | 26 | 0.0000 |

#### Accelerated Failure Time Model Results

Multivariable AFT models of the Weibull, log-logistic, and log-normal distributions were fitted to the time-to-first ANC visit data by taking into account all the covariates that were significant at the 25% level of significance in the univariable analysis. The covariate’s place of residence, spouses’ levels of education, birth order, religion, family wealth index, media access, mother’s involvement in healthcare decisions, and distance to a medical facility were all significant in the final model according to Table 4. The covariate mother’s work status was significant in the log-logistic and log-normal models but not in the Weibull model. Age group was not significant in any AFT model at the 5% level of significance.

**Table 4.** Result of the final Multivariable log-normal Accelerated Failure Time model.

| **Variables** | **Category** | **Coef($\beta$)** | **$\Phi$** | **SE($\beta$)** | **p-value** | **95% CI for (□)** |
| --- | --- | --- | --- | --- | --- | --- |
| **Residence** | Urban | Ref |  |  |  |  |
|  | Rural | 0.242 | 1.274 | 0.027 | <0.001 | [1.207 1.344] |
| **Mother's education** | No education | Ref |  |  |  |  |
|  | Primary | -0.163 | 0.850 | 0.023 | <0.001 | [0.812,0.889] |
|  | Secondary & above | -0.207 | 0.813 | 0.037 | <0.001 | [0.757,0.873] |
| **Women's work status** | No | Ref |  |  |  |  |
|  | Yes | -0.050 | 0.951 | 0.018 | 0.004 | [0.918,0.984] |
| **Birth order** | 1 | Ref |  |  |  |  |
|  | 2-3 | 0.045 | 1.046 | 0.024 | 0.062 | [0.998,1.096] |
|  | 4-5 | 0.061 | 1.063 | 0.027 | 0.025 | [1.007,1.121] |
|  | 6+ | 0.179 | 1.196 | 0.028 | <0.001 | [1.133,1.263] |
| **Religion** | Orthodox | Ref |  |  |  |  |
|  | Protestant | 0.206 | 1.229 | 0.025 | <0.001 | [1.170,1.290] |
|  | Muslim | 0.112 | 1.118 | 0.020 | <0.001 | [1.076,1.162] |
|  | Others | 0.459 | 1.583 | 0.078 | <0.001 | [1.359,1.844] |
| **Wealth index** | Poor | Ref |  |  |  |  |
|  | Middle | -0.165 | 0.848 | 0.026 | <0.001 | [0.806,0.892] |
|  | Rich | -0.178 | 0.837 | 0.025 | <0.001 | [0.797,0.879] |
| **Media access** | No | Ref |  |  |  |  |
|  | Yes | -0.134 | 0.874 | 0.021 | <0.001 | [0.838,0.912] |
| **Decision on health care** | No | Ref |  |  |  |  |
|  | Yes | -0.075 | 0.928 | 0.023 | <0.001 | [0.887, 0.970] |
| **Distance** | Not a big problem | Ref |  |  |  |  |
|  | A big problem | 0.128 | 1.137 | 0.018 | <0.001 | [1.096, 1.178] |
| **Husband education** | No education | Ref |  |  |  |  |
|  | Primary | -0.158 | 0.854 | 0.021 | <0.001 | [0.819, 0.891] |
|  | Secondary & above | -0.200 | 0.819 | 0.031 | <0.001 | [0.771, 0.869] |
| **Region** | Tigray | Ref |  |  |  |  |
|  | Afar | 0.499 | 1.646 | 0.045 | <0.001 | [1.506 1.800] |
|  | Amhara | 0.246 | 1.279 | 0.035 | <0.001 | [1.193 1.370] |
|  | Oromia | 0.505 | 1.656 | 0.038 | <0.001 | [1.536 1.785] |
|  | Somali | 0.577 | 1.780 | 0.044 | <0.001 | [1.632 1.942] |
|  | Benishangul | 0.274 | 1.316 | 0.041 | <0.001 | [1.214 1.426] |
|  | SNNPR | 0.264 | 1.302 | 0.040 | <0.001 | [1.202 1.409] |
|  | Gambela | 0.334 | 1.397 | 0.047 | <0.001 | **[1.274 1.532]** |
|  | Harari | 0.241 | 1.273 | 0.047 | <0.001 | **[1.161 1.396]** |
|  | Addis Ababa | 0.186 | 1.205 | 0.047 | <0.001 | **[1.099 1.320]** |
|  | Dire Dawa | -0.066 | 0.937 | 0.048 | 0.171 | **[0.853 1.029]** |
| **Scale parameter ($\lambda$) = 0.652** |  | **log-Likelihood = - 6538.73** |  |  | **AIC = 13135.46** |  |

#### Model comparison for Accelerated Failure Time models

Using the factors that are significant in multivariable analysis, a model comparison was conducted. The AIC was used to compare the efficacy of various models. It is the most widely used factor that may be used to choose the ideal model. The AIC for Weibull, Log-logistics, Log-normal is 13500.61, 13142.22, 13135.46 respectively and the BIC for Weibull, Log-logistics, Log-normal is 13693.17, 13341.66, 13334.9 respectively. The time-to-first ANC visit dataset indicate that the log-normal AFT model (AIC= 13135.46) fit the data the best.

#### Multivariable Analysis Accelerated Failure Time Model

Table 4 below displays the final log-normal AFT model’s output. Extended time to first ANC visit is indicated by an accelerated factor more than one (□>1), whereas a shorter time to first ANC visit is indicated by an accelerated factor less than one (□<1). In comparison to their reference category, the covariates of place of residence, birth order, religion, distance to a health facility, and area exhibited a positive coefficient (□>1), indicating that they had a longer duration until their first ANC visit. The time it takes for pregnant Ethiopian women to receive their first ANC visit is much shortened when mother and husband have higher levels of education, are employed, have families with more money, have access to the media, and participate in health care decisions.

When comparing the acceleration factors (φ) for the Tigray region to those for Afar, Amhara, Oromia, Somali, Benishangul, SNNPR, Gambela, Harari, and Addis Ababa, the results show that all of those women had longer times until their first ANC visit than the Tigray region. For women from the Dire Dawa region, however, the difference is not statistically significant (p=0.171).

#### Gamma Shared Frailty Model

Gamma shared frailty model with three baseline distribution were fitted by using region as a frailty term and it is highly significant for all the three baseline distributions whereas it was not significant in inverse Gaussian shared frailty distribution using Weibull and log logistic baseline. The log-logistic gamma shared frailty model had a smaller AIC (12947.93) than Weibull (13521.46) and Log-normal (13018.05) gamma shared frailty model. The result also showed that the value of the shared frailty (θ) is 0.127, 0.527 and 0.157 for Weibull, log-logistic and Log-normal gamma shared frailty models respectively; the heterogeneity between regions was high when estimated by log-logistic gamma shared frailty model, which was 0.527. The Kendall’s tau (τ) is higher for higher values of theta (θ) which measure the association within region. The estimated τ = 0.209 shows that there is strong dependence within the cluster (region). This indicates log-logistic gamma shared frailty model is more efficient model to describe time-to-first ANC visit dataset.

The result in Table 5 shows that the frailty in this model is assumed to follow a gamma distribution with mean 1 and variance equal to theta (θ). The estimated value of theta (θ) is 0.527. A likelihood ratio test for the hypothesis θ =0 is shown in the last row of Table 5 indicate a chi-square value of 620.42 with one degree of freedom resulted a highly significant p-value of <0.001. This implied that the frailty component had significant contribution to the model. And the associated Kendall’s tau (τ), which measures dependence within clusters (region), is estimated to be 0. 209. The estimated value of the shape parameter (γ) in this model is 11.273. This value showed the shape of hazarded function is Unimodal because the value is greater than unity it increases up to optimum point and then decreases.

**Table 5.** Results of the final multivariable log-logistics gamma shared frailty model.

| Variables | Category | | Coef($\beta$) | SE($\beta$) | $\Phi$ | p-value | 95% CI | for (□) |
| --- | --- | --- | --- | --- | --- | --- | --- | --- |
| Residence | Urban |  | Ref |  |  |  |  |  |
|  | Rural |  | 0.255 | 0.040 | 1.291 | <0.001 | [1.193, 1.397] |  |
| Mothers educational level | No education |  | Ref |  |  |  |  |  |
|  | Primary |  | -0.097 | 0.022 | 0.908 | <0.001 | [0.870, 0.947] |  |
|  | Secondary& above |  | -0.139 | 0.033 | 0.870 | <0.001 | [0.815, 0.929] |  |
| Birth order | 1 |  | Ref |  |  |  |  |  |
|  | 2-3 |  | 0.062 | 0.022 | 1.064 | 0.0051 | [1.019, 1.112] |  |
|  | 4-5 |  | 0.075 | 0.026 | 1.077 | 0.0038 | [1.024, 1.133] |  |
|  | 6+ |  | 0.189 | 0.027 | 1.208 | <0.001 | [1.147, 1.273] |  |
| Religion | Orthodox |  | Ref |  |  |  |  |  |
|  | Protestant |  | 0.035 | 0.028 | 1.036 | 0.2131 | [0.980, 1.095] |  |
|  | Muslim |  | -0.126 | 0.023 | 0.882 | <0.001 | [0.843, 0.922] |  |
|  | Others |  | 0.329 | 0.081 | 1.389 | <0.001 | [1.186, 1.628] |  |
| Wealth index | Poor |  | Ref |  |  |  |  |  |
|  | Middle |  | -0.131 | 0.025 | 0.878 | <0.001 | [0.835, 0.922] |  |
|  | Rich |  | -0.143 | 0.024 | 0.867 | <0.001 | [0.827, 0.909] |  |
| Media access | no |  | Ref |  |  |  |  |  |
|  | ye |  | -0.117 | 0.023 | 0.890 | <0.001 | [0.851, 0.930] |  |
| Decision on health care | No |  | Ref |  |  |  |  |  |
|  | Yes |  | -0.046 | 0.022 | 0.955 | 0.0414 | [0.914, 0.998] |  |
| Distance to health facility | Not a big problem |  | Ref |  |  |  |  |  |
|  | A big problem |  | 0.076 | 0.018 | 1.079 | <0.001 | [1.041, 1.117] |  |
| Husbands educational level | No education |  | Ref |  |  |  |  |  |
|  | Primary |  | -0.090 | 0.021 | 0.914 | <0.001 | [0.878, 0.952] |  |
|  | Secondary& above |  | -0.168 | 0.029 | 0.845 | <0.001 | [0.799, 0.895] |  |
| Residence with media access | Urban | No | Ref |  |  |  |  |  |
|  | Rural | Yes | 0.156 | 0.046 | 1.169 | 0.007 | [1.068, 1.280] |  |

$\lambda = 0.292$ $\gamma = 11.273$ log- Likelihood = - 6453.965
Theta ( $\theta$ ) = 0.527 Kendall's ( $\tau$ ) = 0. 209 AIC = 12947.93
Likelihood-ratio test of $\theta = 0$ : chi-square2(01) = 620.42, Prob>=chibar2 = 0.000\*
Ref. Reference, Coef: Coefficient of parameter, SE ( $\beta^{\wedge}$ ): standard error for $\beta^{\wedge}$ ; $\phi$ = Acceleration factor; 95%CI $\phi$ : 95% confidence interval for ( $\phi$ ); $\theta$ = frailty variance; $\lambda$ = scale parameter; $\gamma$ = Shape parameter

The result in Table 5 shows that the confidence intervals of the accelerated factor for all significant categorical covariates do not include one at 5% level of significance. This shows that they are significant factors for determining the survival of time to first ANC visit among pregnant women in Ethiopia. However, from the covariate mother’s Protestant religion is not significant by taking orthodox religion as reference (p-value = 0.2131, □= 1.036, 95% CI, (0.980, 1.095). The covariate place of residence, birth order, protestant religion and distance to health facility had acceleration factor greater than one (□>1), indicating this covariate extending time to first ANC visit, while education level of spouses, wealth index of the family, women’s media access, mother’s involvement on health care decision and Muslim religion had acceleration factor less than one (□<1) indicating this covariate shortened time to first ANC visit compared with to the reference category.

#### Comparison of Log-normal AFT and Log-logistic Gamma shared Frailty model

When the mother who was not educated was used as a reference, the acceleration factor (□) for women who attended primary, secondary, and higher education levels was 0.908 and 0.870 with 95% confidence intervals (0.870, 0.947) and (0.815, 0.929) respectively, at the 5% significance level. This shows that compared to an illiterate mother, an educated mother had a shorter survival period until her first ANC visit.

With one birth order used as a reference category, the acceleration factor (□) and its 95% confidence interval for the mother’s birth orders of 2-3, 4-5, and 6+ were, respectively, 1.064 (1.019, 1.112), 1.077 (1.024, 1.133), and 1.208 (1.147, 1.273). Mothers with two or more birth orders had a later first ANC visit than mothers with one birth order, and mothers with six or more birth orders had a longer wait time to the first ANC visit—by a factor of 1.207 at the 5% level of significance than mothers with one birth order. These accelerated factors were greater than one.

With the poor as the reference, the accelerated factor (□) and its 95% confidence interval (CI) for the wealth index of the middle-class and rich families were, respectively, 0.878 (0.835 0.922) and 8657 (0.827, 0.909). A significant p-value of less than 0.001 indicates that the family’s wealth index was a statistically significant factor for the survival of the time until the first ANC visit.

The mother who had access to the media had an acceleration factor (□) of 0.890 with a 95% confidence interval (CI) of (0.851, 0.930); the fact that υ is less than one indicates that the time to the first ANC visit was shorter for women who used media access than for those who did not. Compared to women who did not participate in health care choice-making (no decision as reference; 95% CI: (0.914, 0.998)), moms who participated in health care decision-making reduced the duration to the first ANC visit by a factor of 0.955.

Using a non-educated husband as a reference, the acceleration factor (□) for women whose husbands had only completed primary school was estimated to be 0.914 with a 95% confidence interval (CI) of (0.878, 0.952) and 0.845 (8.749) for secondary education and above. Compared to women whose husbands were illiterate, moms whose husbands were educated had shorter survival times for time to first ANC visit by a factor of 0.914 and 0.952, respectively.

At the 5% significance level, the distance to the health facility had a statistically significant effect on the time until a pregnant mother in Ethiopia had her first ANC visit (p-value<0.001). For the mother’s long-distance trip to the medical facility, the acceleration factor (υ) was 1.079 with a 95% confidence interval (1.041, 1.117). This shows that mothers who travel a considerable distance to a health facility have a longer time until their first ANC visit by a factor of 1.079 at the 5% significance level than mothers who reside close to a health facility.

Using orthodox religion as a reference, the acceleration factor (□) for mothers who follow the Muslim faith was estimated to be 0.882 with 95% CI (0.843, 0.922) while for mothers who follow other religions, it was 1.389 with 95% CI (1.186, 1.628). This suggests that, even while there is no appreciable difference in the duration to the first ANC visit between women’s protestant and orthodox religious followers, Muslim religious followers had a shorter survival time than orthodox religious followers.

The women who lived in rural areas and had access to media had an acceleration factor (ω) of 1.169 with a 95% confidence interval (CI) of 1.068 to 1.280. This indicates that the women who had media access and lived in rural areas had a longer survival time before their first ANC visit compared to the women who lived in urban areas and had no media access.

## Model diagnostics

### Cox-Snell residual plots

One method for determining how well the model fits the data is to look at the Cox-Snell residuals. Fig 3 below shows the plot for the fitted model of residuals for log logistic to our data using cumulative hazard function and maximum likelihood estimate. The cumulative hazard function against Cox-Snell residuals graphic indicates that the log logistic baseline distribution is nearly a straight line through the origin with slop 1, indicating that it is more suited for the time to first ANC visit data set than the Weibull and log normal distributions.

## Discussion

The main aim of the study was to assess the timing of pregnant women’s first antenatal care visit and to identify its determinant factors among pregnant mothers in Ethiopia using AFT and parametric shared frailty models by considering three baseline distributions (Weibull, log-logistic, log-normal distributions) and gamma frailty.

According to WHO recommendation on ANC for a positive pregnancy experience; every pregnant mother should start ANC booking during the first trimester of pregnancy (WHO, 2018). However, in this study out of the total 7559 pregnant women in Ethiopia, only one-fifths (20.4%) of mothers started their first ANC visit within the right time, and the rest 79.6% started late. This is consistent with studies conducted in Ethiopia Gamo Gofa [14], Dembecha [15] 6%, Wollega [14], 19%, 21.7% respectively and Nigeria [16] 17.4%; but this finding is comparatively lower than the study finding from Ethiopia Addis Ababa [17] 42%, Gondar [18], 35%, Debre Markos [9] 66.6% and Tigray [8] 41% and from abroad Tanzania [19] 29%.

The estimated median survival time of the first ANC visit during pregnancy of Ethiopia pregnant women is found to be 18 weeks (4.5 months) with 95% confidence interval (4.1, 4.9). This is almost analogous to a previous study from Ethiopia which was 4.5 months from Gondar [18], 5 months in Tigray [8] and 5 months in Tanzania [19]. This finding is almost similar with the EDHS-2016 report of 4.7 months [7]. This study result is relatively lower compared to the finding of another study reported 6 months in Nigeria [16].

Multivariable analysis using log-logistic gamma shared frailty model show that from predisposing factors, place of residence, women’s educational level, birth order, religion, media access, women’s involvement on health care decision, husband’s educational level and from enabling factors wealth index of the family and distance to health facility were significantly associated with time to first ANC visit during pregnancy in Ethiopia at 5% level of significant. The result of the current review is in agreement with the systematic review and meta-analysis by Gezahegn Tesfaye [20] and other studies conducted in Ethiopia, Tigray [8], Gondar [18], Wollega [21], Denbecha [15], in Tanzania [19] and in Nigeria [16]. But gender of the household head, women’s marital status, women’s work status, pregnancy type and age of the mother are not significant predictor of timing of first ANC visit.

The results of this study suggested that place of residences is significant predictive factor for tome-to-first ANC visits of Ethiopian pregnant women. Those pregnant women’s living in rural area was 1.291 times late to start their first ANC visit as compared to their counterpart with urban area. The finding is supported by the study conducted in Ethiopia East Wollega zone, [20], Gondar Town [18], Tigray [8], and abroad Ethiopia in Tanzania [19] and Nigeria [16].

This study showed that women’s education level has a significant effect on timing of first ANC visit, indicates that women with primary, secondary and above education level start ANC visit earlier than illiterate women. This result agreed with those studies conducted in Ethiopia, East Wollega zone [20], North West Gondar Town [18], Tigray [8]. It could be reasonable to say that educated women, compared to uneducated, have better access to information, possess a level of health literacy that could empower them to exercise their choice and overcome cultural barriers of ANC service utilization [22].

The finding of this study revealed that religion had a significant effect on the time to first ANC visit. This shows that there was the difference in the timing of ANC visits along religion, where women who followed the Muslim religion had a shorter time to first ANC visit. The study conducted in the East Wollega zone, Ethiopia [20] and Nigeria [16] shows the same result.

As part of enabling factors, this study shows that the economic status of the family is a significant determinant factor of the timing of first ANC visit. Pregnant women who had high household income shortened the time to first ANC visit by a factor of 0.867 times as compared to those households with low economy. This finding is in line with studies conducted in Ethiopia, East Wollega Zone [20], North West Gondar Town [18], Tigray Ethiopia [8], and abroad Ethiopia in Tanzania [19] and Nigeria [16]. This is because the fact that those mothers in wealthy households used the health service more than those in poor and middle-income ones, high income increases the ability to pay for health care services, transportation, and other indirect costs and better education.

Access to media is one of the determinant factors of time to the first ANC visit. The findings of this study show that those pregnant women who exposed to media have 0.890 times shortened time to their first ANC visit as compared to their counterparts who had no access to media. The finding is supported by the study conducted in the Central Zone, Tigray Ethiopia [8], south Ethiopia [23], and East Wollega Zone [20].

The result of this study shows that women’s involvement in health care decisions is another variable found that is associated with the timing of the first ANC visit. Women who had been involved in health care decisions shortened their first ANC visit by a factor of 0.955 times than those who had not been involved. This could be because noninvolved women were under the influence of their family or husband (especially in male-headed householder families), restricted from complying with family norms, and had a lack of family or social support. The finding is supported by the study done in the East Wollega zone, Ethiopia [20], North West Gondar Town Ethiopia [18], Tanzania [19] and Nigeria [16].

In this study as part of enabling factors revealed that distance to health facilities was found to be a determinant factor of the first ANC visit. Pregnant women who lived far from the health facility were 1.079 times more likely to be started late for their first ANC visit as compared to their counterparts who lived near to health facility. The finding is supported by the study conducted in Nigeria [16], women who live within an hour’s walking distance from the health facility were about eight times more likely to visit ANC than those above an hour distance walking.

This study also found that women’s having husband who attended primary, secondary, and above education level start ANC visits earlier than those women whose husbands never attended formal education. This result agreed with those studies conducted in Ethiopia, East Wollega zone [20], and Tigray Ethiopia [8]. Education of the mother and husband could play a great role in improving awareness of health matters and the importance of ANC in particular, having awareness enables women to seek ANC and utilize the service early in pregnancy [20].

This study finds that the gender of the household head, women’s marital status, women’s work status, pregnancy type, and age of the mother are not significant predictors of the timing of the first ANC visit. which is in line with the prior study conducted in Nigeria [16] and Ethiopia, [20], [21]. A study from Nigeria [16] also shows that the variable pregnancy type does not affect timing of the first ANC visit. Other studies in Tigray Ethiopia (Gidey et al., 2017) and East Wolloga (Ejeta et al, 2017) find that mothers’ age is not significantly associated with the time to first ANC visit. In contrast to this study result, studies from Nigeria [16], Tanzania [19], Gondar [18], and Wollega [20], revealed that mother’s age and women’s work status, were significantly associated with first ANC visit. Mothers aged greater than 25 years and below were more likely to commence at the recommended time compared to their counterparts [23]; and employed women were less likely to delay their ANC as compared to their counterparts [20]; a study from Tanzania [19] and Gondar Ethiopia [18], shows that unmarried younger women were more likely to booking ANC visit early than married women. This difference may be due to sample size, time of study, and population composition.

## Conclusions

The study subjects (pregnant women) in this study came from clustered communities and hence clustered survival data correlated at the regional level, indicates the presence of heterogeneity and necessitates between regions; assuming that women within the same region share similar unobserved risk factors. The results of the log-logistic gamma shared frailty model showed that place of residence, mother’s and husband’s level of education, birth order, religion, wealth index, media access, women’s involvement in health care decisions and distance to health facility were found significant predictors for time-to-first ANC visit of pregnant women in Ethiopia. In general, this study illustrates that women’s and their husbands primary and high level of education, having media access, urban resident women, involved on health care decision, orthodox and Muslim religion follower, women’s who had lower birth order, lived near to health facility and increasing wealth index of the family were significantly shorten time-to-first ANC visit, while women’s residing in rural areas, other religion follower, had higher birth order, had no media access, not involved on health care decision and women’s lived far from health facility have significantly prolong time to first ANC visit. From the category of women’s religion, protestant was not statistically significant. The current finding illustrates that highest proportion of women in Ethiopia still late for first ANC booking, which in turn implies that many women are at risk of several obstetric complication.

## Contributors

Conceptualization: Yitagesu Eshetu, Zelalem Tolosa, Tigist Getachew

Data curation: Yitagesu Eshetu, Tigist Getachew

Formal analysis: Yitagesu Eshetu, Zelalem Tolosa, Tigist Getachew

Investigation: Yitagesu Eshetu

Methodology: Yitagesu Eshetu, Zelalem Tolosa, Tigist Getachew

Project administration: Yitagesu Eshetu

Resources: Yitagesu Eshetu, Tigist Getachew

Software: Yitagesu Eshetu, Zelalem Tolosa, Tigist Getachew

Supervision: Zelalem Tolosa

Writing original draft: Yitagesu Eshetu

Writing review & editing: Yitagesu Eshetu, Tigist Getachew

## Funding

The authors have not declared a specific grant for this research from any funding agency in the public, commercial or not-for-profit sectors.

## Competing interests

None declared.

## Patient and public involvement

Patients and/or the public were not involved in the design, or conduct, or reporting, or dissemination plans of this research.

## Patient consent for publication

Not applicable.

## Ethics approval

The study was based on a secondary data analysis of the EDHS data, and it is publicly available. We obtained permission from MEASURE DHS/ICF International and used the data. During the original data collection of DHS data, international and national ethical guidelines were applied. Additionally, the study participants gave written consent during data collection. Provenance and peer review Not commissioned; externally peer reviewed.

## Data availability statement

All data relevant to the study are included in the article or uploaded as supplementary information. The survey datasets used in this study were based on a publicly available dataset that is freely available online with no participant’s identity from http://www.dhsprogram.com/data/available-datasets. cfm. Approval was sought from MEASURE DHS/ICF International, and permission was granted for this use.

## Notes

### Competing Interest Statement

The authors have declared no competing interest.

### Funding Statement

The author(s) received no specific funding for this work.

## References

[1] WHO, “5. Antenatal care 29,” pp. 29–54, 2018.

[2] A. Program, “Pnadi869,” 2005.

[3] W. B. G. and U. N. P. D. WHO, UNICEF, UNFPA, “Trends in Maternal Mortality : 1990 to 2015: Estimates Developed by WHO,UNICEF,UNFPA, The World Bank and the United Nations Population Divisions,” Who /Rhr/15.23, vol. 32, no. 5, pp. 1–55, 2015, [Online]. Available: http://whqlibdoc.who.int/publications/2010/9789241500265_eng.pdf

[4] H. Blencowe et al., “National, regional, and worldwide estimates of stillbirth rates in 2015, with trends from 2000: A systematic analysis,” Lancet Glob. Heal., vol. 4, no. 2, pp. e98–e108, 2016, doi: 10.1016/S2214-109X(15)00275-2.

[5] S. Neupane, B. I. Nwaru, Z. Wu, and E. Hemminki, “Work behaviour during pregnancy in rural China in 2009,” Eur. J. Public Health, vol. 24, no. 1, pp. 170–175, 2013, doi: 10.1093/eurpub/ckt135.

[6] J. Karlsson, “Child Marriage a Mapping of Programmes and Partners in Twelve Countries in East and Southern Africa,” pp. 1–82, 2017.

[7] E. Central Statistic Agency, Ethiopian Demographic and Health survey Final Report. 2016.

[8] G. Gidey, B. Hailu, K. Nigus, T. Hailu, W. Gher, and H. Gerensea, “Timing of first focused antenatal care booking and associated factors among pregnant mothers who attend antenatal care in Central Zone, Tigray, Ethiopia,” BMC Res. Notes, vol. 10, no. 1, pp. 1–6, 2017, doi: 10.1186/s13104-017-2938-5.

[9] A. A. Ewunetie, A. M. Munea, B. T. Meselu, M. M. Simeneh, and B. T. Meteku, “DELAY on first antenatal care visit and its associated factors among pregnant women in public health facilities of Debre Markos town, North West Ethiopia,” BMC Pregnancy Childbirth, vol. 18, no. 1, pp. 1–8, 2018, doi: 10.1186/s12884-018-1748-7.

[10] M. Carpenter, Survival Analysis: A Self-Learning Text, vol. 39, no. 2. 1997. doi: 10.1080/00401706.1997.10485091.

[11] D. R. Cox, “cox_jrssB_1972_hi_res.pdf.” 1972.

[12] R. G. Gutierrez, “Parametric Frailty and Shared Frailty Survival Models,” Stata J. Promot. Commun. Stat. Stata, vol. 2, no. 1, pp. 22–44, 2002, doi: 10.1177/1536867x0200200102.

[13] D. Collet, Modelling Survival Data in Medical Research. Fourth edition. | Boca Raton□: Taylor and Francis, 2023. |Series: Chapman & Hall/CRC texts in statistical science. 2023. [Online]. Available: https://www.routledge.com/

[14] F. Gebremeskel, Y. Dibaba, and B. Admassu, “Timing of First Antenatal Care Attendance and Associated Factors among Pregnant Women in Arba Minch Town and Arba Minch District, Gamo Gofa Zone, South Ethiopia,” J. Environ. Public Health, vol. 2015, 2015, doi: 10.1155/2015/971506.

[15] M. Gedefaw, B. Muche, and M. Aychiluhem, “Current Status of Antenatal Care Utilization in the Context of Data Conflict: The Case of Dembecha District, Northwest Ethiopia,” Open J. Epidemiol., vol. 04, no. 04, pp. 208–216, 2014, doi: 10.4236/ojepi.2014.44027.

[16] A. F. Fagbamigbe, B. Mashabe, L. Lepetu, and C. Abel, “Are the timings and risk factors changing? Survival analysis of timing of first antenatal care visit among pregnant women in Nigeria (2003-2013),” Int. J. Womens. Health, vol. 9, pp. 807–819, 2017, doi: 10.2147/IJWH.S138329.

[17] A. Tariku, Y. Melkamu, and Z. Kebede, “Previous utilization of service does not improve timely booking in antenatal care: Cross sectional study on timing of antenatal care booking at public health facilities in Addis Ababa,” Ethiop. J. Heal. Dev., vol. 24, no. 3, pp. 226–233, 2010, doi: 10.4314/ejhd.v24i3.68390.

[18] T. W. Gudayu, M. Solomon, and A. Abdella, “Timing and factors associated with first antenatal care booking among pregnant mothers in Gondar,” BMC Pregnancy Childbirth, vol. 14, no. 287, pp. 5–7, 2014.

[19] K. Gross, “Intermittent Preventive Treatment during Pregnancy and Antenatal Care in Practice: A study from the Kilombero Valley, Tanzania,” p. 188, 2012, [Online]. Available: https://edoc.unibas.ch/19025/1/Intermittent_Preventive_Treatment_during_Pregnancy_and_Anten.pdf

[20] G. Tesfaye, D. Loxton, C. Chojenta, A. Semahegn, and R. Smith, “Delayed initiation of antenatal care and associated factors in Ethiopia: A systematic review and meta-analysis,” Reprod. Health, vol. 14, no. 1, 2017, doi: 10.1186/s12978-017-0412-4.

[21] E. Ejeta, R. Dabsu, O. Zewdie, and E. Merdassa, “Factors determining late antenatal care booking and the content of care among pregnant mother attending antenatal care services in east wollega administrative zone, west Ethiopia,” Pan Afr. Med. J., vol. 27, pp. 1–7, 2017, doi: 10.11604/pamj.2017.27.184.10926.

[22] E. S. Greenaway, J. Leon, and D. P. Baker, “Understanding the association between maternal education and use of health services in ghana: Exploring the role of health knowledge,” J. Biosoc. Sci., vol. 44, no. 6, pp. 733–747, 2012, doi: 10.1017/S0021932012000041.

[23] M. B. Geta and W. W. Yallew, “Early Initiation of Antenatal Care and Factors Associated with Early Antenatal Care Initiation at Health Facilities in Southern Ethiopia,” Adv. Public Heal., vol. 2017, pp. 1–6, 2017, doi: 10.1155/2017/1624245.

